# Comparing Expert and Computerised Pattern Identification in Antepartum Cardiotocography

**DOI:** 10.1101/2025.08.15.25333416

**Authors:** John Tolladay, Beth Albert, William R. Cooke, Manu Vatish, Gabriel Davis Jones

## Abstract

**Objective:** To assess the reliability of antepartum cardiotocography (CTG) pattern identification by quantifying the level of agreement among expert clinicians and between clinicians and the Dawes–Redman (DR) computerised system, particularly in the context of limited formal guidelines and the known subjectivity of antepartum assessments. The findings are intended to support improvements in training and the standardization of CTG interpretation.

**Methods:** Five senior clinicians with expertise in fetal monitoring independently annotated 105 15-minute fetal heart rate (FHR) traces using structured web-based software. For each trace, participants identified the baseline, accelerations and decelerations and categorised variability. Inter-observer agreement was assessed using intraclass correlation (ICC) for baselines and Fleiss’ *κ* for variability. Sensitivity and positive predictive value (PPV) for acceleration and deceleration detection were calculated relative to majority-voted results. The DR algorithm was used to identify the same patterns and the output was compared against the clinical consensus annotations.

**Results:** Baseline agreement was excellent among participants (ICC = 1.0, 95% CI 1.00– 1.00). Variability classifications showed only moderate concordance (Fleiss’ *κ* = 0.39, 95% CI *−*0.01–0.56). Detection of accelerations and decelerations varied across clinicians (sensitivity 39.2–97.2%, PPV 39.7–91.3%). The DR system showed good agreement for accelerations (sensitivity = 64.8%, 95% CI 57.1–72.1%; PPV = 85.0%, 95% CI 79.5–89.8%) but poor agreement for decelerations (sensitivity = 50.0%, 95% CI 14.3–75.0%; PPV = 20.0%, 95% CI 4.2–39.1%). DR-classified variability showed minimal agreement with clinical ratings (Fleiss’ *κ* = 0.002, 95% CI *−*0.007–0.027).

**Conclusions:** Antepartum CTG interpretation remains inconsistent for identification of decelerations and variability. While baseline assessment appears robust, current clinical and algorithmic approaches show limited agreement for more ambiguous patterns. These findings support the need for updated training and refined algorithms to improve reliability in antepartum fetal surveillance.

## 1. Introduction

Fetal monitoring plays a central role in antenatal care. Cardiotocography (CTG), which records the fetal heart rate (FHR) and uterine activity, is one of the most commonly used methods to assess fetal well-being before birth. Despite its widespread use, human visual CTG interpretation remains highly subjective, with significant inter-observer variability even among experienced clinicians [1]. In the intrapartum setting, inter- and intra-observer agreement between experts is estimated to be as low as 29%, and false-positive rates for detecting fetal risk are as high as 60% [2, 3, 4, 5, 6, 7]. This has important, clinical implications, triggering avoidable interventions or late identification of fetal compromise [8, 9, 10, 11].

Expert clinical evaluation of intrapartum CTG may fail to identify 35–92% of FHR patterns [12, 13]. Classification systems, such as those proposed by FIGO[14] and NICE[15] aim to standardise intrapartum interpretation and improve agreement in the identification of important CTG patterns [16]. However, such efforts have focused almost entirely on intrapartum monitoring. By contrast, antepartum CTG (despite its critical differences in clinical context and physiology) remains under-investigated and no equivalent national or international guidelines exist. A small number of earlier studies explored inter-observer agreement for antepartum CTG interpretation, but these were conducted decades prior, do not reflect current clinical practice or the widespread adoption of digital CTG and automated analysis systems [12, 13, 17, 18]. The reliability of human interpretation in this setting is poorly understood. This poses a significant concern, given the routine use of antepartum monitoring and the potential for misinterpretation to precipitate adverse maternal and fetal outcomes.

The Dawes-Redman (DR) system is one of the most widely adopted tool for computerised antepartum CTG analysis. By applying numerical algorithms to the FHR signal, it evaluates 10 predefined criteria to assess the condition of the fetus [19, 20]. Meeting all criteria indicates fetal well-being, while referral for expert clinical assessment is recommended if they are not met within 60 minutes. The DR system is incorporated into both national and local guidelines and has become the standard in many settings [19, 21, 22]. Although highly effective for standard CTG assessment, its sensitivity to subtle but clinically important FHR patterns (e.g., atypical decelerations or subtle variability changes) is not well established [23]. Its performance relative to expert visual pattern recognition has also not been rigorously examined.

This study evaluates the consistency of expert identification of antepartum CTG patterns and compares the results with outputs from the DR system, focusing on the identification of patterns indicative of fetal well-being or distress. Comparison will assess the reliability of the DR system and the potential for automated systems to enhance consistency and accuracy. Given the risks of misinterpretation, such as missed fetal distress or unnecessary intervention, enhancing the reliability of CTG analysis is crucial. Discrepancies between human and automated pattern recognition also requires further investigation to clarify clinical significance and the implications for patient care.

## 2. Methods

### 2.1 Data Curation and Expert Annotation

A total of 105 15-minute cardiotocography (CTG) traces (1,575 minutes) were selected by a panel comprising a midwife and medical doctor, each with expertise in CTG interpretation. Eligible traces were required to be free from corruption or missing data and to contain one or more of the following features: accelerations, decelerations, periods of low, medium or high variability, or the absence of any such features. Traces were sampled exclusively from the antepartum period, as the Dawes-Redman system is designed for use in this setting, and because intrapartum CTGs differ markedly in both physiology and clinical purpose. Clinical metadata such as gestational age, fetal sex or indication for monitoring were not provided to participants. This was intentional: currently, no established guidelines stratify CTG interpretation by gestational age, sex, or clinical indication in the antepartum setting. Furthermore, to evaluate pattern recognition objectively, we aimed to assess how reliably CTG features could be identified without the influence of contextual information or pregnancy outcomes. In clinical practice, CTG features should be recognised objectively and only subsequently integrated with patient history and risk factors. The uterine activity trace (toco) was not included, in keeping with standard practice for antepartum CTG interpretation, where uterine contractions are not routinely assessed.

FHR annotations were performed using a web-based application, adapted from a previously published tool to enable the capture of additional CTG patterns and participant characteristics [24]. No identifiable participant information was accessible to the study team. Each participant was presented with 15-minute FHR segments from the selected CTG dataset. Traces were displayed using the standard UK aspect ratio (1 cm/min), with grid lines marking every 30 seconds on the x-axis and every 20 bpm on the y-axis. Maternal-reported fetal movements were indicated by blue triangles superimposed on the display. For each FHR segment, participants placed markers along the trace to indicate the estimated baseline at that time-point. Markers were then automatically interpolated to generate a continuous baseline. Accelerations and decelerations were annotated by drawing boxes from the onset to the end of each pattern. FHR variability was categorized as “low”, “medium” or “high” based on visual assessment. Participants were able to revise annotations prior to final submission. An illustration of the annotation interface is provided in the supplementary material (Figure S1).

### 2.2 Participant Recruitment and Guidance

Five clinicians - three midwives and two consultant obstetricians - were recruited from the clinical staff at the John Radcliffe Hospital, Oxford, UK. The most experienced available personnel were selected to ensure an expert-level analysis of the CTG segments. All participants were previously trained in both the antepartum and intrapartum CTG interpretation and had more than 10 years of clinical experience, with routine exposure to CTGs. They were blinded to the results of any automated or previous analysis performed on the CTG traces. All participants were provided standardised instructions at the start of their session, consistent with the annotation procedures described previously. Participants were requested to assess variability using thresholds applied in “fresh eyes” reviews [25]. This involves visually estimating whether the variability is *<* 5 bpm (“reduced”), *>* 25 bpm (“saltatory”) or within these two limits (“normal”).

Participants were instructed that completed segments could not be revisited, to minimise bias and ensure that identification of patterns in one trace did not influence the annotation of subsequent segments. The application was presented at a fixed size on a standardised display for all participants. Study team members were available at the beginning of each session to provide guidance on the annotation process, after which participants completed the task independently, with assistance available only for non-interpretive issues.

### 2.3. Data Processing

For each annotated segment, the start and end times of accelerations and decelerations were recorded. Baseline values were saved as a time series of FHR values in beats per minute and variability was stored as a categorical variable. CTG segments were then analysed using the Dawes-Redman (DR) system for comparison with participant annotations. Variability was classified using the same thresholds as the participants, based on the long-term variability calculated by the system for each segment. Baselines and acceleration/deceleration start and end points were extracted directly from the DR analysis results.

In the absence of specific antepartum guidelines, annotations were performed according to FIGO criteria, chosen for their familiarity to participants and similarity in specification of FHR patterns[14]. The application was configured to restrict acceleration and deceleration annotations to a minimum duration of 15 seconds. Patterns with a peak deviation from the average baseline over the same interval of less than 15 bpm were excluded during postprocessing. Equivalent restrictions were placed on the output of the DR system to ensure comparable results.

Majority voting was used to derive consensus annotations. For any time period in which more than half of the participants identified a pattern, the average start and end times across those annotations were used to define a single representative interval. Overlapping patterns of the same type were merged in to a single reading. Baselines were calculated as the median of all participant’s baselines at each time-point. For variability, the mode of the participants’ selections was taken as the consensus classification.

### 2.4. Statistical Analysis

Inter-observer agreement was assessed using the intraclass correlation coefficient (ICC) for baseline identification and Fleiss’ *κ* for variability classification. To evaluate concordance with the majority-voted results, accelerations and decelerations annotated by participants and detected by the DR system were assessed using sensitivity and positive predictive value (PPV). A detected pattern was classified as a true positive if it met one of the following three criteria:

1. The detected pattern was entirely contained within the majority-voted reference interval.
2. One boundary (start or end) of the detected pattern fell within the reference interval, and the opposite boundary extended no more than 60 seconds beyond the reference boundary.
3. The reference interval was entirely contained within the detected pattern, with no more than 60 seconds excess duration beyond the reference bounds.

Patterns that did not meet these criteria were classified as false positives or false negatives, depending on whether they were present in the evaluated set or the referenced standard. Confidence intervals were estimated using bootstrapping. For Fleiss’ *κ*, inter-observer agreement was assessed by resampling participants with replacement. Sensitivities and PPVs were calculated by resampling CTG segments with replacement and evaluating the start and end point agreement within the resampled set.

## 3. Results

**Table 1:**
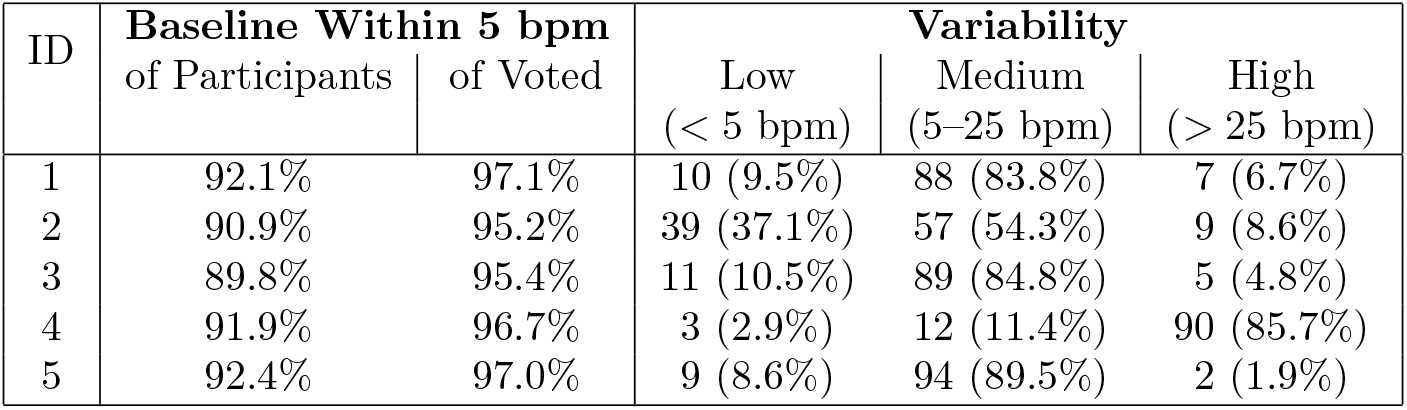
Inter-observer agreement on baseline estimation and variability classification across participants. Baseline agreement is shown as the proportion of baseline points within ±5 bpm between each participant and all others (“Within 5 bpm of Participants”), and relative to the majority-voted median baseline (“Within 5 bpm of Voted”). All values exceeded 89%, indicating high consistency in baseline identification. Variability classifications are presented as counts and percentages of segments rated as “low” (< 5 bpm), “medium” (5–25 bpm), or “high” (> 25 bpm) by each participant. While most segments were classified as “medium,” systematic differences were observed across raters, particularly in the use of “low” and “high” categories, reflecting inter-observer subjectivity in variability assessment.

| ID | Baseline Within 5 bpm |  | Variability |  |  |
| --- | --- | --- | --- | --- | --- |
| | of Participants | of Voted | Low<br>($< 5$ bpm) | Medium<br>(5–25 bpm) | High<br>($> 25$ bpm) |
| 1 | 92.1% | 97.1% | 10 (9.5%) | 88 (83.8%) | 7 (6.7%) |
| 2 | 90.9% | 95.2% | 39 (37.1%) | 57 (54.3%) | 9 (8.6%) |
| 3 | 89.8% | 95.4% | 11 (10.5%) | 89 (84.8%) | 5 (4.8%) |
| 4 | 91.9% | 96.7% | 3 (2.9%) | 12 (11.4%) | 90 (85.7%) |
| 5 | 92.4% | 97.0% | 9 (8.6%) | 94 (89.5%) | 2 (1.9%) |

### 3.1. Baseline Assignment

There was strong agreement among participants in baseline assignment (Table 2). Participant 3 showed the lowest concordance, though still high, with 89.8% of their baseline points falling within ±5 bpm of the others. Overall agreement was excellent, with 91.4% of baseline points within ±5 bpm across all participants and an intraclass correlation coefficient of 1.0 (95% CI 1.0–1.0). The largest discrepancies occurred in segments with shifting baselines or high variability without sustained periods of stability (Figure 1 for examples).

**Table 2:** Detection performance and annotation exclusions for accelerations and decelerations by individual expert clinicians. Each participant independently annotated accelerations and decelerations across 105 antepartum CTG segments. The total number of marked patterns (“Count”) and the number excluded for failing to meet FIGO thresholds for amplitude and duration are shown. Sensitivity and positive predictive value (PPV) were calculated by comparing each participant’s annotations to the majority-voted reference standard, with 95% confidence intervals. Exclusion rates ranged from 16.7% to 25.3% for accelerations and from 20.3% to 42.4% for decelerations. Sensitivity varied substantially between participants for both features, particularly for decelerations, where sensitivity ranged from 45.7% to 97.2% and PPV from 39.7% to 91.3%.

| ID | Accelerations |  |  |  |
| --- | --- | --- | --- | --- |
|  | Count | Excluded | Sensitivity (%) | PPV (%) |
| 1 | 324 | 54 (16.7%) | 85.1 (80.7–89.9) | 76.3 (68.4–82.9) |
| 2 | 126 | 23 (18.3%) | 39.2 (28.7–49.5) | 91.3 (83.5–97.2) |
| 3 | 479 | 108 (22.5%) | 86.0 (80.5–90.7) | 63.1 (55.9–70.1) |
| 4 | 324 | 82 (25.3%) | 86.6 (80.9–92.1) | 88.4 (82.7–93.1) |
| 5 | 335 | 83 (24.8%) | 87.7 (83.3–92.0) | 84.9 (79.1–90.2) |

Table 2: Detection performance and annotation exclusions for accelerations and decelerations by individual expert clinicians.
| ID | Decelerations |  |  |  |
| --- | --- | --- | --- | --- |
|  | Count | Excluded | Sensitivity | PPV |
| 1 | 56 | 14 (25.0%) | 75.7 (58.6–88.2) | 66.7 (47.6–82.5) |
| 2 | 118 | 50 (42.4%) | 77.1 (60.0–94.5) | 39.7 (25.9–54.2) |
| 3 | 74 | 15 (20.3%) | 84.2 (70.0–93.8) | 54.2 (41.3–68.1) |
| 4 | 58 | 13 (22.4%) | 97.2 (90.6–100.0) | 77.8 (62.5–91.9) |
| 5 | 28 | 7 (25.0%) | 45.7 (28.6–67.7) | 76.2 (58.8–95.2) |

**Figure 1.**
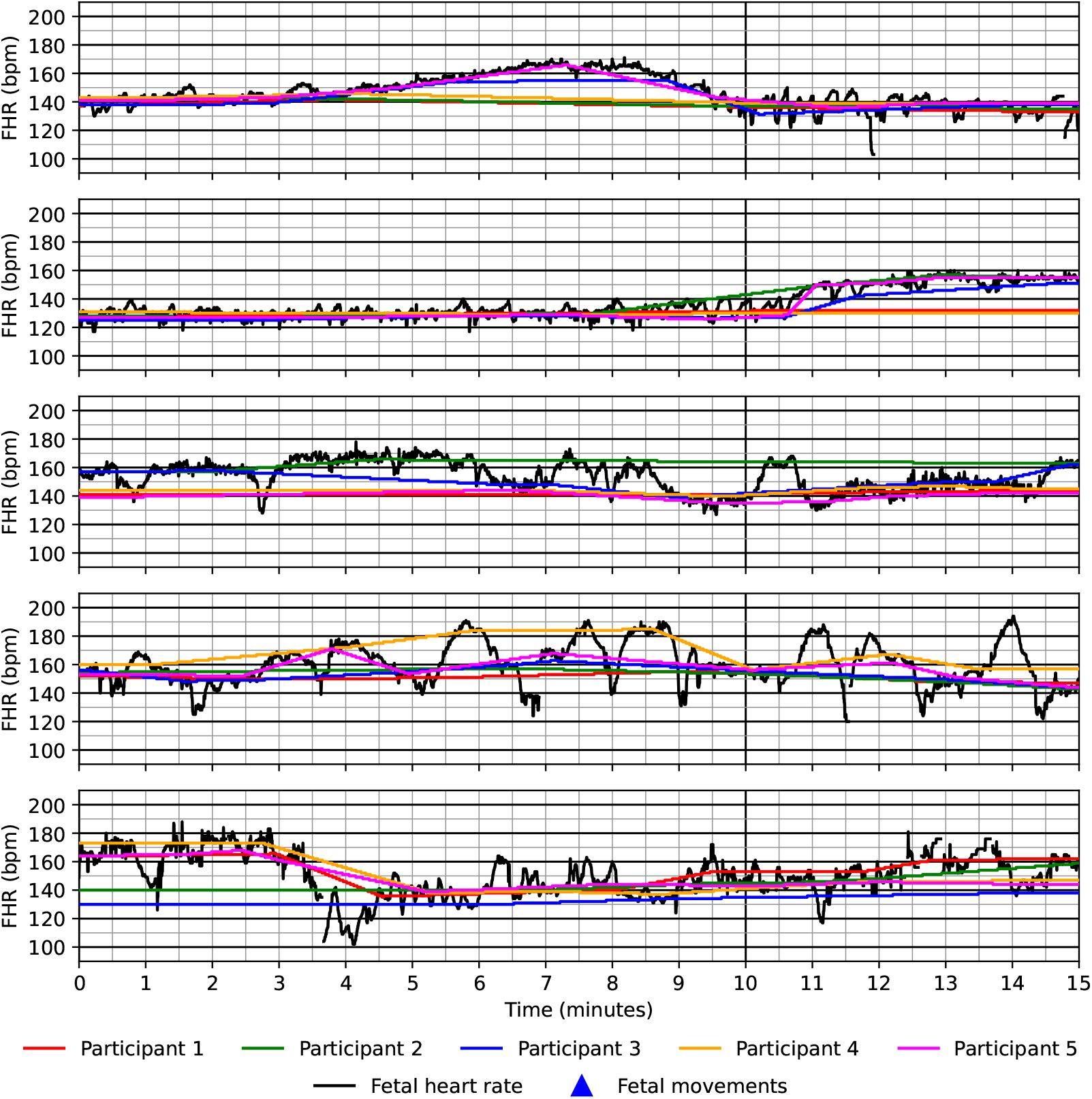
Baseline annotations from five clinicians for fetal heart rate segments with the greatest interobserver discrepancy. Each panel displays a 15-minute fetal heart rate (FHR) segment overlaid with the baseline annotations from five expert participants. Segments were selected based on the largest differences in baseline placement across annotators. Despite high overall agreement (91.4% of values within *±*5 bpm), discrepancies emerged in segments with unstable or shifting baselines, where signal variability complicated visual estimation. Coloured lines represent individual participant baselines, and black traces represent the FHR signal. Triangles indicate maternally reported fetal movements. Axes are not displayed using the standard CTG aspect ratio; the y-axis has been stretched to enhance visibility of inter-rater differences.

### 3.2. Variability Classification

Most CTG segments were classified as having “medium” variability. Participant 2 assigned “low” variability to 29 (27.6%) more segments than any other participant, whereas participant 4 more frequently selected “high” variability, applying this classification to 90 segments (85.7%). Despite these tendencies, overall agreement on variability was moderate (Fleiss’ *κ* = 0.39, 95% CI *−*0.01–0.56).

### 3.3. Acceleration and Deceleration Identification

Across the five participants, 1,588 accelerations and 334 decelerations were collectively annotated (2). For failing to meet FIGO criteria for amplitude and duration [14], 350 accelerations (22.0%) and 99 decelerations (29.6%) were excluded. Two of the participants (IDs 1 and 2) showed a greater tendency to over-identify decelerations, with 25.0% and 42.4% of their annotations excluded, respectively, compared with 16.7% and 18.3% of their accelerations. Most of the excluded patterns (74.6%) were within 5 bpm of the FIGO threshold for amplitude.

Figure 2 illustrates an example segment showing accelerations and decelerations as annotated by individual participants, the majority-voted consensus, and the Dawes-Redman system. The first deceleration identified by DR was also marked by two of the five participants, whereas the second was annotated by four participants and included in the majority-voted result. One participant annotated a continuous acceleration between 11 and 13 minutes. However, most participants interpreted a dip within this interval as a boundary, resulting in two separate accelerations in the consensus annotation. A deceleration near the 14-minute mark was identified by three participants but not detected by the DR system. The figure also includes one acceleration and one deceleration that were excluded due to insufficient magnitude. Both were borderline cases and their classification depends on the baseline assigned by each participant (Figure 3).

**Figure 2.**
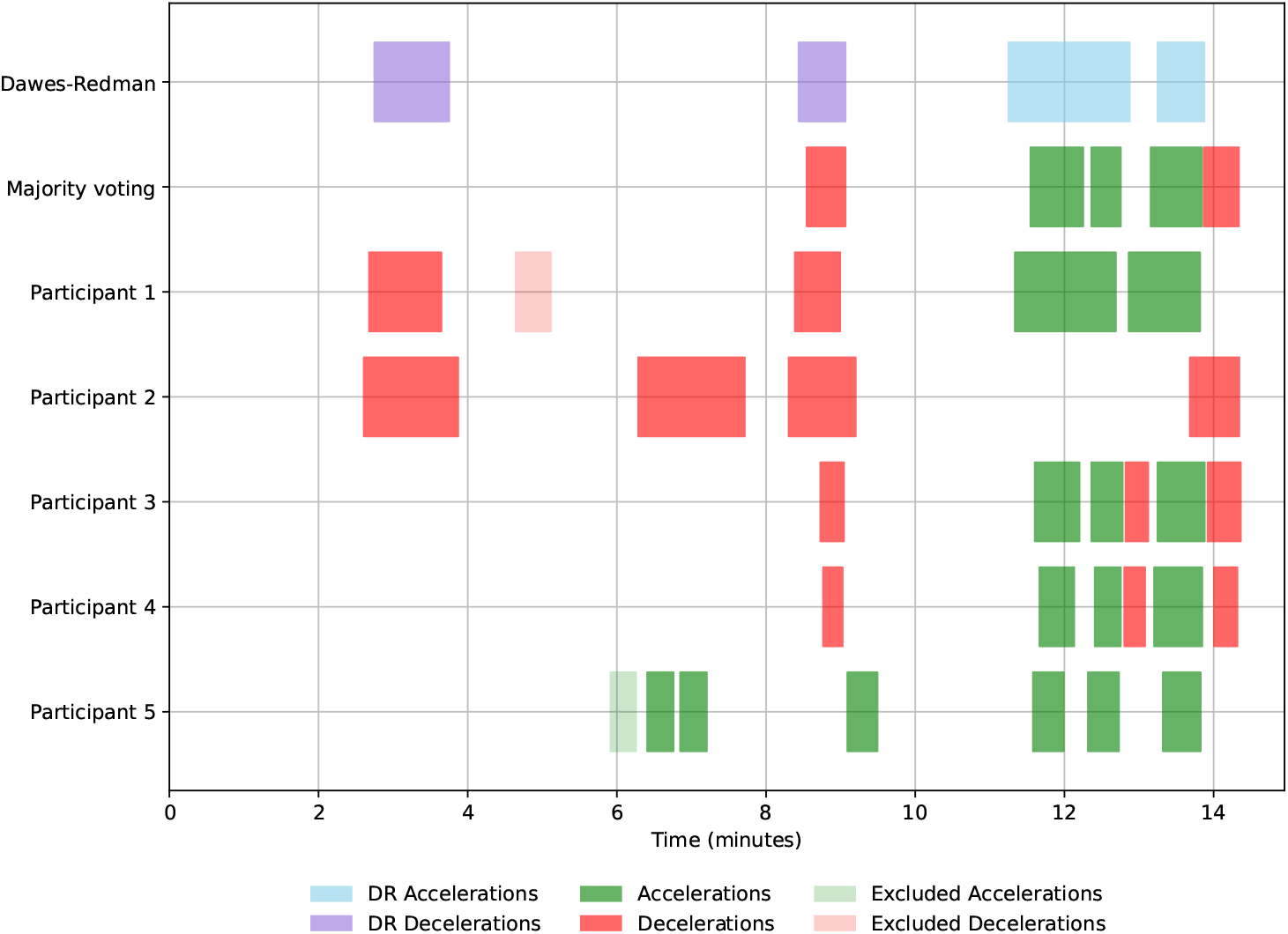
Comparison of pattern annotations by clinicians and the Dawes-Redman system for a single 15-minute fetal heart rate segment. This example illustrates the timing and classification of accelerations and decelerations as identified by five senior clinicians, majority voting, and the Dawes-Redman (DR) system. Green and red boxes represent accelerations and decelerations, respectively, as annotated by individual clinicians. Shaded boxes denote patterns excluded post hoc for failing to meet FIGO criteria (minimum 15 bpm amplitude and 15 second duration). Blue and purple boxes indicate accelerations and decelerations detected by the DR system. Areas of concordance and discordance between DR and clinician annotations are visible, including DR-identified patterns with limited expert agreement and a deceleration marked by the majority of participants but missed by DR. This demonstrates areas of agreement and divergence in pattern recognition across human and automated interpretation.

**Figure 3.**
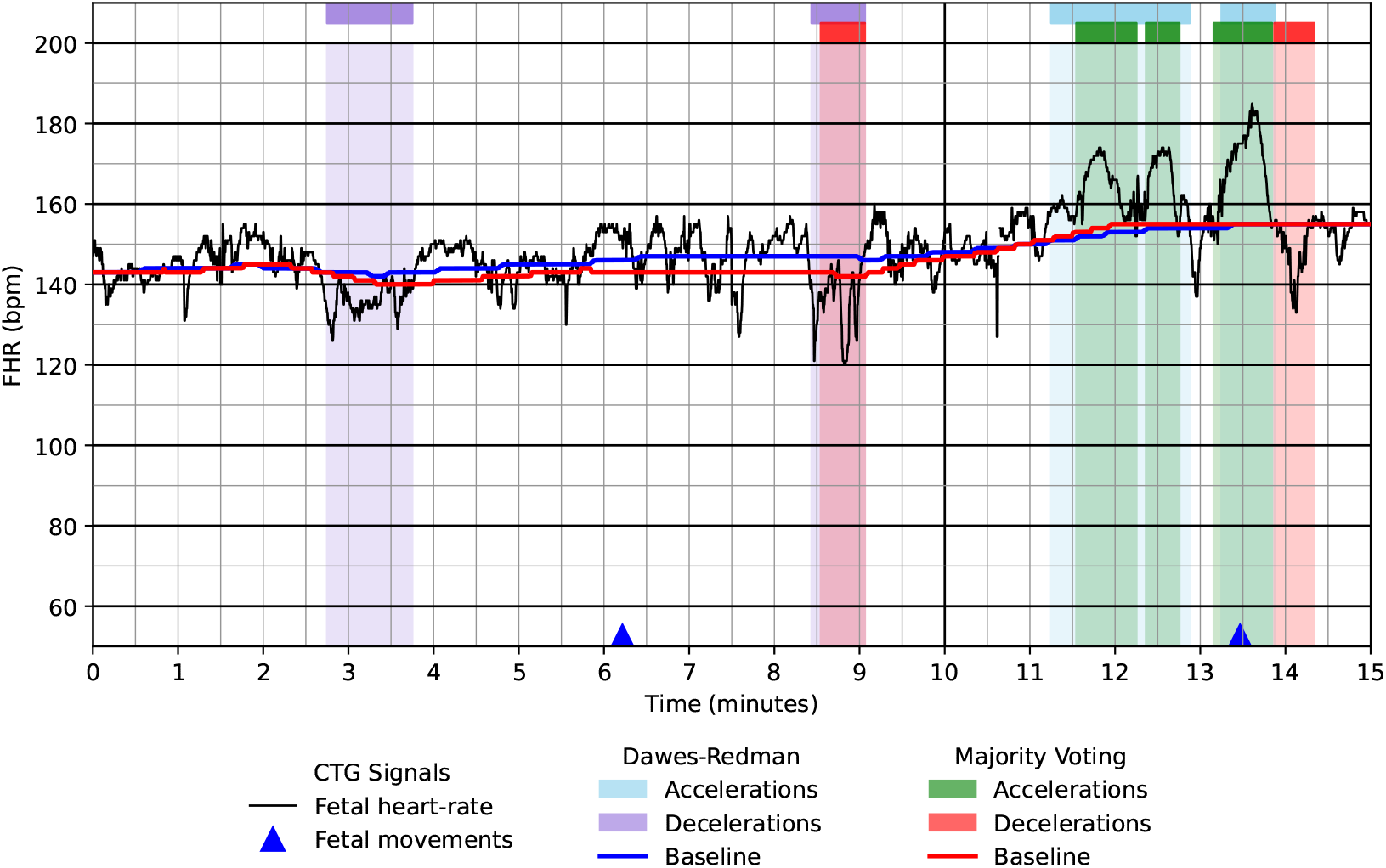
Comparison of baseline assignment and event detection by the Dawes-Redman system and majority-voted clinician annotations for a single fetal heart rate (FHR) segment. A 15-minute FHR trace is shown with superimposed annotations from the Dawes-Redman (DR) system and majority-voted expert clinician interpretations. The black line represents the raw FHR signal and blue triangles indicate maternally reported fetal movements. Shaded regions correspond to accelerations (green) and decelerations (red) as identified by majority vote, and to DR-identified accelerations (blue) and decelerations (purple). The baseline determined by each method is plotted in red (majority vote) and blue (DR). This segment was classified as having “medium” variability by both DR and most participants. Several differences in event detection are visible, including one deceleration marked by DR but not by clinicians, and a set of accelerations split by clinicians into two separate events, while DR identified a single continuous episode.

When compared to the majority-voted annotations, participant sensitivity for detecting accelerations ranged from 39.2–87.7% (mean = 76.9%, 95% CI 50.7–100.0%) and for decelerations from 45.7–97.2% (mean = 76.0%, 95% CI 52.5–99.5%). Overall, participants demonstrated good concordance with the consensus, though individual variability was observed. For example, participant 5 showed a 42.0% difference for sensitivity between acceleration detection (87.7%, 95% CI 83.3–92.0%) and deceleration detection (45.7%, 95% CI 28.6–67.7%). Participants 2 and 3 tended to over-identify decelerations, yielding lower PPV (39.7%, 95% CI 25.9–54.2%, and 54.2%, 95% CI 41.3–68.1%, respectively) despite relatively high sensitivities (77.1%, 95% CI 60.0–94.5%, and 84.2%, 95% CI 70.0–93.8%). In contrast, participant 2 under-identified accelerations, with markedly lower sensitivity (39.2%, 95% CI 28.7–49.5%) than PPV (91.3%, 95% CI 83.5–97.2%).

### 3.4. Comparison to Dawes-Redman System

The DR system demonstrated moderate agreement with majority-voted annotations for accelerations, with a sensitivity 64.8% (95% CI 57.1–72.1%) and PPV of 85.0% (95% CI 79.5– 89.8%) (Table 3). Agreement for decelerations was substantially lower, with a sensitivity of 50.0% (95% CI 14.3–75.0%) and a PPV of 20.0% (95% CI 4.2–39.1%).

**Table 3:** Agreement between the Dawes-Redman system and majority-voted expert annotations. Performance of the Dawes-Redman (DR) system in identifying accelerations and decelerations was evaluated against a majority-voted reference standard derived from five expert clinicians. Baseline agreement was assessed using the intraclass correlation coefficient (ICC), and agreement on variability classification using Fleiss’ *κ*. Sensitivity and positive predictive value (PPV) are presented with 95% confidence intervals. The DR system showed high concordance with expert-assigned baselines (ICC = 1.00, 90.2% within *±*5 bpm) and good performance in detecting accelerations (sensitivity 64.8%, PPV 85.0%). Performance for deceleration detection was lower (sensitivity 50.0%, PPV 20.0%). Variability classification showed negligible agreement with clinical ratings (Fleiss’ *κ* = 0.002), with the system classifying 93.3% of traces as “medium” variability and none as “high.”

| Accelerations |  |
| --- | --- |
| Sensitivity (%) | 64.8 (57.1–72.1) |
| PPV (%) | 85.0 (79.5–89.8) |
| Decelerations |  |
| Sensitivity (%) | 50.0 (14.3–75.0) |
| PPV (%) | 20.0 (4.2–39.1) |
| Baseline |  |
| ICC | 1.00 (1.00–1.00) |
| Within 5 bpm (%) | 90.2% |
| Variability |  |
| Fleiss' $\kappa$ | 0.002 (–0.007–0.027) |
| Low (< 5 bpm) | 7 (6.7%) |
| Medium (5–25 bpm) | 98 (93.3%) |
| High (> 25 bpm) | 0 (0.0%) |

Figure 3 illustrates key differences between automated and clinician-annotated pattern detection. The first deceleration identified by the DR system was not annotated by the majority of participants, possibly because the magnitude was close to the FIGO threshold. Whether the pattern was marked depended on each participant’s assigned baseline and visual judgement of amplitude. Between 11 and 13 minutes, the DR system annotated a single acceleration, whereas most participants identified a brief trough that separated the interval into two distinct accelerations in the majority-voted result. A deceleration near the 14-minute mark was annotated by multiple participants but was not detected by the DR system, potentially due to its close temporal proximity to a preceding large acceleration.

Baseline agreement between the DR system and participants remained high (ICC = 1.00, 95% CI 1.00–1.00), with 90.2% of values falling within ±5 bpm. In contrast, agreement on variability classification was lower for the DR system (Fleiss’ *κ* = 0.002, 95% CI *−*0.007– 0.027), compared with moderate agreement between participants (Fleiss’ *κ* = 0.39, 95% CI *−*0.01–0.56). The DR system classified 98 of 105 segments as having “medium” variability (5–25 bpm), with only 7 classified as “low” (*<* 5 bpm) and none as “high” (*>* 25 bpm), suggesting a strong bias toward mid-range values and limited dynamic range in variability detection.

## 4. Discussion

Despite widespread use in antenatal care, antepartum cardiotocography (CTG) remains under-studied compared to intrapartum CTG. Research and guidelines focus almost exclusively on intrapartum monitoring, with limited investigation into clinical antepartum interpretation or concordance with computerised systems [14]. This gap is striking given the global reliance on antepartum CTG for fetal assessment and its role in determining pregnancy trajectories [26]. This study addresses that gap by systematically comparing expert identification of FHR patterns with the Dawes-Redman (DR) system, providing the first detailed, segment-level analysis of inter-observer agreement of feature-level annotation and algorithmic concordance in antepartum CTG interpretation.

### 4.1. Human Visual Interpretation

Decelerations were the pattern identified with the least consistency among clinicians (sensitivity 45.7–97.2%; PPV 39.7–77.8%). Participants frequently included patterns with amplitudes near the FIGO threshold (15 bpm), indicating a tendency to consider borderline cases clinically relevant regardless of formal criteria. This underscores the subjectivity of visual interpretation and the need for careful standardisation. These variations are clinically meaningful, as decelerations can be associated with fetal hypoxia, placental insufficiency or umbilical cord compression [21, 27]. Missed decelerations may delay escalation and risk poor perinatal outcomes, whereas false positives can lead to avoidable interventions like iatrogenic preterm birth or unnecessary induction.

Conversely, baseline identification was highly reliable, with 91.4% of baseline points within ±5 bpm. This suggests that baseline assessment is a robust component of CTG interpretation. For variable baselines, clinicians differed in whether they followed the upper or lower limits of the FHR trend. This can influence whether fluctuations are interpreted as accelerations or decelerations, contributing further to interpretive variability.

Variability classification showed greater subjectivity than baseline or acceleration detection. Inter-observer agreement was fair to moderate, with some participants demonstrating systematic preferences for particular classifications, but most disagreement occurring between adjacent categories (e.g., “low” vs “medium”). Despite this, thresholds were applied with internal consistency, suggesting standardisation could reduce variability of interpretation. Low FHR variability is associated with adverse outcomes, including fetal acidaemia, hypoxic-ischaemic encephalopathy and stillbirth [19, 20, 28]. Identification of reduced variability is essential for risk stratification, as both over- and under-calling can influence clinical decisions, potentially leading to unnecessary intervention or missed fetal compromise.

These findings support the need for more quantitative approaches to support variability assessment. Current FIGO definitions rely on visual estimation, but integrating numeric metrics (e.g., short- and long-term variability) into clinical displays may reduce subjectivity and enhance early identification of fetal compromise [14].

Importantly, clinicians demonstrated a high degree of engagement and precision. Most patterns (particularly accelerations and baselines) were consistently identified across observers, emphasizing the value of clinical judgement in CTG interpretation. The structured format and experienced experts used in this study helped reduce noise and provided a credible consensus framework.

### 4.2. Computerised Interpretation by the Dawes-Redman System

The DR system demonstrated excellent concordance with clinician-assigned baselines and performed reasonably well for acceleration detection (sensitivity 64.8%; PPV 85.0%). This supports its utility as a standardised baseline estimator and an acceptable tool for identifying reassuring features such as accelerations.

However, performance was more limited for patterns associated with potential pathology. The DR system detected only 50.0% of decelerations identified by most clinicians and showed low PPV (20.0%), often missing subtle or borderline patterns. This could result in under-recognition of emerging compromise, especially in high-risk pregnancies. While strict thresholds reduce false positives, they may also limit detection of early or atypical signs of fetal distress.

Similarly, agreement with clinicians on variability classification was negligible, with 93.3% of segments labelled as “medium”. This narrow distribution suggests limited sensitivity to clinically relevant variation within observed segments and highlights limitations of automated, rule-based categorisation in a domain where subtle dynamics may be significant.

These limitations should be weighed against the system’s consistency, reproducibility and widespread adoption. Unlike human interpretation, it eliminates inter-observer variability and has been associated with reductions in unnecessary admissions and improved workflow standardisation. The DR system also extends beyond pattern recognition to generate a comprehensive assessment of fetal well-being [19, 20]. In a recent study of over 3,300 antepartum traces, the DR system ruled out adverse outcomes with high specificity (90.7%) and achieved a particularly high negative predictive value (99.1%) in low-risk settings, supporting its role in reassuring clinicians and avoiding over-intervention [23]. This broader evaluative capacity helps mitigate limitations observed when analysing individual features in isolation, such as decelerations or variability classifications.

### 4.3. Clinical Implications

This study identifies deceleration detection and variability classification as the principal sources of interpretive disagreement in antepartum CTG. These patterns are central to clinical decision-making, particularly in cases of reduced fetal movements, maternal comorbidities or prolonged gestation. Misclassification may result in missed hypoxia or unnecessary intervention, each carrying significant risk.

A lack of standardised antepartum guidelines, variation in clinical training and differences in CTG display format can contribute to inconsistent interpretation. Standardising training and displays across devices (e.g., bedside monitors, printed outputs, digital tools) may reduce this variability, particularly in resource-limited or decentralised settings where support tools can reduce reliance on highly trained specialists.

For borderline or uncertain features, a hybrid approach may offer added value. Semiautomated systems could allow clinicians to include or exclude specific patterns and observe how these affect the system’s overall risk assessment. This would support exploration of diagnostic uncertainty, combining algorithmic consistency with clinical judgement, improving interpretive accuracy and confidence in decision-making.

### 4.4. Comparison with Intrapartum Studies

Our findings reveal patterns of inter-observer variability consistent with those observed in intrapartum settings. Clinicians demonstrate higher agreement on well-defined patterns like baseline and accelerations, while variability assessment and overall classification remain more subjective. For instance, studies using the 2015 FIGO guidelines reported good interobserver proportion of agreement (Pa) for baselines (Pa = 0.85) and decelerations (Pa = 0.92) but lower agreement for variability (Pa = 0.82) and overall CTG classification (Pa = 0.60) [29]. Similarly, midwives’ intrapartum CTG interpretations showed fair to good agreement (*κ* = 0.65–0.74), with lowest agreement found for baseline variability (*κ* = 0.50) [30]. These parallels suggest that subjective visual analysis challenges persist across settings, underscoring the need for enhanced standardisation and training in both antepartum and intrapartum CTG interpretation.

### 4.5. Strengths and Limitations

This study’s strengths include in its focus on antepartum CTG – a relatively unexplored area – and its use of structured expert annotations to generate a high-quality reference dataset. All participants had advanced training and over 10 years CTG interpretation experience, ensuring clinical relevance. The standardised interface and fixed display settings reduced visual variability, while the majority-voting provided a reference standard for future algorithm benchmarking and development. Although the panel size was limited, interobserver agreement studies commonly use small expert panels. When participants are highly experienced and the annotation process is structured, such studies can still yield robust insights into clinical variability [16]. Participants were drawn from a single academic centre in the UK, which may affect generalisability.

## 5. Conclusion

This study is the first to evaluate expert interpretation of antepartum CTG at the pattern level and compare it directly with output from the Dawes-Redman system. Agreement among clinicians was high for baseline identification but notably lower for deceleration detection and variability classification features critical to identification of fetal compromise. The Dawes-Redman system aligned well with clinicians for accelerations and baselines but demonstrated limited sensitivity and poor positive predictive value for decelerations, and classified nearly all traces as “medium” variability. These findings suggest that while baseline interpretation is robust, both human and algorithmic approaches struggle with features more indicative of fetal pathology. Improved training, standardised displays and refinement of algorithmic logic (particularly for decelerations and variability) are needed to enhance consistency and clinical reliability in antepartum CTG interpretation.

## Data Availability

All data produced in the present study are available upon reasonable request to the authors.

## 6. Author Contributions

John Tolladay: Conceptualisation, data curation, formal analysis, investigation, methodology, software, resources, validation, visualisation, writing - original draft, writing - review & editing. Beth Albert: Conceptualisation, methodology, project administration, writing - review & editing. William R. Cooke: Validation, writing - review & editing. Manu Vatish: Validation, writing - review & editing. Gabriel Davis Jones: Conceptualization, formal analysis, funding acquisition, investigation, methodology, project administration, resources, supervision, validation, visualisation, writing - review & editing.

## 7. Declarations

This study was approved by the Ethics Committee in the Joint Research Office, Research and Development Department, Oxford University Hospitals NHS Trust (approval number: 25/HRA/1966, granted on 13th May 2025) and supported by the Medical Research Council (UKRI grant MR/X029689/1). The anonymised data that support the findings of this study are available upon reasonable request from the corresponding author.

