## Supplementary Material for "Comparing Expert and Computerised Pattern Identification in Antepartum Cardiotocography"

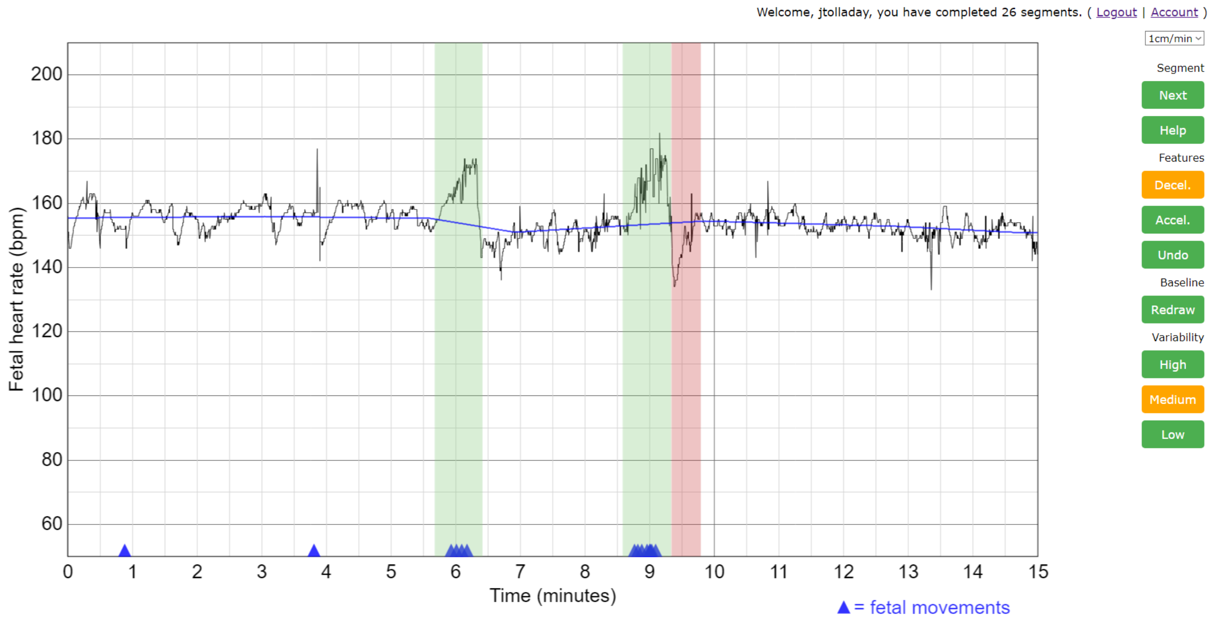


**Figure S1:** An example fetal heart rate segment displayed in the application used by the participants for annotation. The fetal heart rate (black line) and maternal indications of fetal movements (blue triangles) are displayed to the participant. Accelerations (green shading) and decelerations (red shading), and the baseline (blue line) are drawn on to the CTG by the participants. Variability classification is assigned using one of the three buttons on the right-hand side under the heading “Variability”.
